# Understanding national trends in COVID-19 vaccine hesitancy in Canada – April 2020 to March 2021

**DOI:** 10.1101/2021.11.10.21266174

**Authors:** Kim L. Lavoie, Vincent Gosselin-Boucher, Jovana Stojanovic, Samir Gupta, Myriam Gagne, Keven Joyal-Desmarais, Katherine Seguin, Sherri Sheinfield-Gorin, Paula Ribeiro, Brigitte Voisard, Michael Vallis, Kim Corace, Justin Presseau, Simon L. Bacon, for the iCARE Study Team

## Abstract

**Objective:** Key to reducing COVID-19 morbidity and mortality and reducing the need for further lockdown measures in Canada and worldwide is widespread acceptance of COVID-19 vaccines. Vaccine hesitancy has emerged as a key barrier to achieving optimal vaccination rates, for which there is little data among Canadians. This study examined rates of vaccine hesitancy and their correlates among Canadian adults.

**Methods:** This study analyzed data from five age, sex and province-weighted population-based samples to describe rates of hesitancy between April 2020 and March 2021 among Canadians who completed online surveys as part of the iCARE Study, and various sociodemographic, clinical and psychological correlates. Vaccine hesitancy was assessed by asking: “*If a vaccine for COVID-19 were available today, what is the likelihood that you would get vaccinated?*” Responses were dichotomized into ‘very likely’, ‘unlikely’, ‘somewhat unlikely’ (reflecting some degree of vaccine hesitancy) vs ‘extremely likely’ to get the vaccine, which was the comparator.

**Results:** Overall, 15,019 respondents participated in the study. A total of 42.2% of respondents reported vaccine hesitancy over the course of the study, which was lowest during surveys 1 (April 2020) and 5 (March 2021) and highest during survey 3 (November 2020). Fully adjusted multivariate logistic regression analyses revealed that women, those aged 50 and younger, non-Whites, those with high school education or less, and those with annual household incomes below the poverty line in Canada (i.e., $60,000) were significantly more likely to report being vaccine hesitant over the study period, as were essential and healthcare workers, parents of children under the age of 18, and those who do not get regular flu vaccines. Believing engaging in infection prevention behaviours (like vaccination) is important for reducing virus transmission and high COVID-19 health concerns (being infected and infecting others) were associated with 77% and 54% reduction in vaccine hesitancy, respectively, and having high personal financial concerns (worried about job or income loss) was associated with 1.33 times increased odds of vaccine hesitancy.

**Conclusion:** Results point to the importance of targeting vaccine efforts to women, younger people and socioeconomically disadvantaged groups, and that vaccine messaging should emphasize the benefits of getting vaccinated, and how the benefits (particularly to health) far outweigh the risks. Future research is needed to monitor ongoing changes in vaccine intentions and behaviour, as well as to better understand motivators and facilitators of vaccine acceptance, particularly among vulnerable groups.

## Introduction

The SARS-CoV-2 virus causing coronavirus disease (COVID-19) has caused a global pandemic, resulting in significant morbidity, mortality and economic and social disruption in Canada and around the world. Key to reducing disease morbidity and mortality and reducing the need for future lockdowns is widespread acceptance of COVID-19 vaccines, several of which have been approved for those aged 12 and older by Health Canada,^1^ with approvals pending for children aged 5-11. High rates of vaccine acceptance was thought to be necessary for achieving target levels of herd immunity,^2^ but it has proven difficult to estimate the minimum threshold of immunization needed to achieve this due to the emergence of highly virulent strains like [Delta] whose R0 has been estimated to be 5 to 6 times greater than the original Wuhan SARS-CoV-2 strain.^3,4^ This has led experts to recommend vaccinating as much of the population as possible and exploring the need for additional, ‘booster’, or yearly doses.^5^ Regardless of how COVID-19 vaccination schedules unfold over the short and longer-term, the ultimate success of vaccination programs depends on people’s willingness to get vaccinated. However, several reports from nations where vaccines have been widely available indicate that intentions to get a COVID-19 vaccine have been steadily declining (and rates of vaccine hesitancy steadily increasing) since the first pandemic wave. For example, a longitudinal study in the US reported significant declines in the likelihood of getting vaccinated (somewhat or very likely to get vaccinated), from a high of 74% in early April 2020 to a low of 56% by early December 2020.^6^ These declines were observed for both men and women and in all age, racial/ethnic and education subgroups. Similar trends were also observed in Australia, where 31.9% of Australians reported being less willing to get vaccinated between August 2020 and January 2021, and was particularly prevalent among Indigenous populations and those who did not complete high school.^7^ Since then, there have been 175 studies worldwide have been published on vaccine hesitancy through to the end of August 2021, including 21 reporting data from Canada. According to a living systematic review by Crawshaw et al.,^8^ the interquartile range of vaccine hesitancy was 12-24%, with a mean of 17%. Overall, these results raise important questions about vaccine attitudes and intentions among Canadians, whose willingness to get vaccinated now and in the future will be critical for optimizing the success of Canada’s vaccine strategy and our successful transition out of the pandemic.

Key to optimizing vaccination rates is understanding patterns and correlates of hesitancy over time. This will allow us to improve vaccine policy planning, develop targeted interventions, and enhance tailoring of vaccine messaging to vulnerable groups. To this end, we examined rates of vaccine hesitancy and their correlates among Canadians by analyzing data from five age, sex and province-weighted population-based samples who completed online surveys between April 2020 and March 2021. In order to explore the factors associated with vaccine hesitancy over time, data across all surveys were examined as a function of key sociodemographics, clinical characteristics, and psychological factors known to be important for vaccine behaviour.^9^

## Methods

### Study Design

The International COVID-19 Awareness and Responses Evaluation (iCARE) Study (www.icarestudy.com)^10^ is an ongoing, international, multi-wave, cross-sectional observational survey study of public awareness, attitudes, and responses to COVID-19 public health policies. The study is led by researchers from the Montreal Behavioural Medicine Centre (MBMC: www.mbmc-cmcm.ca) in collaboration with a team of over 200 international collaborators from more than 40 countries. The survey was designed with international experts to assess constructs from the Capability, Opportunity, Motivation – Behaviour (COM-B) Model of the Behaviour Change Wheel^11^ and from the Health Belief Model.^12,13^ The survey also includes questions on sociodemographics, physical and mental health conditions, general health behaviours, previous COVID-19 infection, awareness of local government prevention policies, perceptions and attitudes about these policies, adherence to prevention behaviours, COVID-19-related concerns and impacts, and vaccine attitudes and intentions (the survey can be found at: www.osf.io/nswcm). The primary REB approval was obtained from the Comité d’éthique de recherche du Centre intégré universitaire de santé et de services sociaux du Nord-de-l’île- de-Montréal (CIUSSS-NIM), approval # : 2020-2099 / 25-03-2020. Full details about the background and methodology have been published elsewhere.^10^

### Participants

For this study, we report data from five nationally representative online surveys of Canadians aged 18 years and over using a recognized polling firm who recruits participants through their proprietary online panel (Leger Opinion). This panel includes over 400,000 Canadians, the majority of which (61%) were recruited within the past 10 years. Two thirds of the panel were recruited randomly by telephone, with the remainder recruited via publicity and social media. Using data from Statistics Canada, results were weighted within each province according to the sex and age of the respondents. Then, the weight of each province was further adjusted to represent their actual weight within the Canadian federation. Data were collected between April 9^th^ and 20^th^, 2020 (Survey 1), June 4^th^ and 17^th^, 2020 (Survey 2), October 29^th^ and November 11^th^, 2020 (Survey 3), January 27 and February 7th (Survey 4), and March 11^th^ to 29^th^ (Survey 5), respectively, using a self-administered Computer-Assisted Web Interface.

### Assessment of vaccine intentions and hesitancy

To assess vaccine hesitancy, we asked: “*If a vaccine for COVID-19 were available today, what is the likelihood that you would get vaccinated?*” Response options (very unlikely, unlikely, somewhat likely, extremely likely, I don’t know/prefer not to answer) were dichotomized into ‘very unlikely, unlikely, somewhat likely’ to describe those indicating at least some degree of hesitancy, vs. ‘very likely’ to describe those with very high intentions to get vaccinated. A dichotomous outcome was chosen to identify all those who could benefit from intervention, with those responding ‘very likely’ to get vaccinated treated as the comparator/reference.

### Assessment of psychological factors

We assessed two psychological factors that are often important motivators of engaging in protective health behaviours: perceived importance of engaging in infection prevention behaviours, and the nature and extent of people’s COVID-19-related concerns.^12-14^ Perceived importance of engaging in COVID-19 prevention behaviours (including getting vaccinated) was assessed using a single question: *“To what extent do you believe that the measures asked of you by your government or local health authority* ***are important*** *to prevent and/or reduce the spread of COVID-19?*” Response options (Not at all important, not very important, somewhat important, very important, I don’t know/prefer not to answer) were dichotomized into ‘very important’ vs all others.

To assess the concerns people have about the COVID-19 virus and its impacts, individuals were presented with the following prompt: *“Because of COVID-19, I am concerned about…”*. Respondents then had to indicate the extent which they had 10 specific concerns, choosing among ‘not at all’, ‘very little’, ‘somewhat’, ‘to a great extent,’ and ‘I don’t know/prefer not to answer’. To cluster COVID-19-related concerns, we performed a principal component analysis on a polychoric correlation matrix of the 10 variables in the concerns module (ordinal scale, as detailed above), details of which can be found elsewhere.^15^ We observed a three-component structure that included: ‘Health concerns’, ‘personal financial concerns’ and ‘social and economy concerns’. Mean values (M) and standard deviations (SD) for each of the three components are reported as a score out of four, from 1= ‘not at all’ to 4 = ‘to a great extent’. Internal consistency for the components ranged from satisfactory (social/economy concerns α=0.69) to excellent (personal financial concerns α=0.82; health concerns α=0.91) for the individual components.^15^

### Statistical analysis

Several survey questions included an answer ‘I don’t know/I prefer not to answer’ which was recoded as a missing value and analyses were based on complete case records. Descriptive statistics (weighted means, SDs, and proportions) were calculated to describe the sample in terms of demographic characteristics, across all surveys. Univariate analyses were conducted to examine differences in sociodemographic characteristics (weighted proportions) as a function of vaccine hesitancy across the five time points. Three separate multivariable logistic regression models were performed to assess associations between vaccine hesitancy (dependent variable) and participant sociodemographic (i.e., age, sex, ethnicity, education, employment status, annual household income, parental status, worker status, provincial region) and clinical characteristics (i.e., health risk conditions, history of flu vaccine, previous COVID-19 infection) (independent variables: Model 1), vaccine hesitancy (dependant variable) and perceived importance of prevention behaviours (independent variable: Model 2), and vaccine hesitancy (dependent variable) and the nature and extent of the three types of COVID-19-related concerns (independent variables: Model 3). Analyses were conducted across all surveys combined and models were partially (covariates included age, sex, ethnicity, and survey wave) and fully adjusted (covariates included age, sex, ethnicity, survey wave, education, employment status, annual household income, health risk condition, essential worker, healthcare worker, parental status, history of flu vaccine, COVID-19 test result). All variables were selected a-priori based on pre-exiting data.^9^ Analyses were also conducted as a function of time point/survey to examine trends over time, assessed using the Welch test. All statistical tests were two-sided and a p-value < 0.05 was considered as statistically significant. Statistical analysis was performed in SAS, version 9.4.

## Results

### Participant characteristics

Our sample included a total of 15,019 respondents (survey 1, n=3003; survey 2, n=3005; survey 3, n=3005, survey 4 = 3000, and survey 5 = 3006) who completed a survey between April 9^th^ 2020 and March 29^th^ 2021, with a response rate of 16% (survey 4) to 25% (survey 5) for each survey (which is considered acceptable for online panels^16^). Participant characteristics collapsed across all surveys and then as a function of survey round can be found in **Table 1** and **Supplementary Table S1**, respectively. Respondents across all five surveys were 51.6% female (range 18-95 years) with a mean age 48.1 [SD 17.2] years. The majority of the sample were White (80.8%), had a high school or less education (72.3%), and reported total family annual incomes over $60,000 (51.7%). Nearly half (49.7%) reported being currently employed. Just over 44% reported having at least one physician-diagnosed health risk condition (e.g., cardiovascular or lung disease, cancer, diabetes, obesity), and just over a quarter (26%) reported having a physician-diagnosed psychiatric disorder (e.g., depressive or anxiety disorder). About 16% identified as being an essential service worker, just over 4% identified as being a healthcare worker, and 21.5% identified as being parents of children under 18. Approximately 17% of respondents had gotten tested for COVID-19, with nearly 1% reporting testing positive. Only 43% of respondents reported getting a flu vaccine at least 3 times or more over the past five years. In general, compared to census data available through Statistics Canada, participants across all five surveys were well distributed across provincial regions, age groups, employment status and income, and there were equal proportions of men and women. However, those with a graduate or post-graduate degree and people of color were less represented.

**Table 1.** Participant characteristics (weighted proportions)

| All surveys n = 15,019 |  |  |  |
| --- | --- | --- | --- |
| Variable | N (%) |  |  |
| Sex |  |  |  |
| Male | 7239 | (48.4) |  |
| Female | 7724 | (51.6) |  |
| Age (Numerical) | 48.1 | ± 17.2 |  |
| Age (Categorical) |  |  |  |
| ≤ 25 years | 1808 | (12.2) |  |
| 26-50 years | 6138 | (41.4) |  |
| ≥ 51 | 6897 | (46.5) |  |
| Race/Ethnicity |  |  |  |
| Non-White | 2687 | (18.2) |  |
| White | 12047 | (81.8) |  |
| Education level |  |  |  |
| High school or lower | 10642 | (72.3) |  |
| Graduate or Postgraduate degree | 4085 | (27.7) |  |
| Current employment status |  |  |  |
| Unemployed | 7412 | (50.3) |  |
| Employed | 7338 | (49.7) |  |
| Annual household income |  |  |  |
| <\$60,000/year | 6405 | (48.3) | |
| ≥\$60,000/year | 6853 | (51.7) | |
| Provincial region |  |  |  |
| Western<sup>1</sup> | 4702 | (31.3) |  |
| Ontario | 5762 | (38.4) |  |
| Quebec | 3523 | (23.5) |  |
| Atlantic<sup>2</sup> | 1032 | (6.9) |  |
| High-risk health condition<sup>3</sup> |  |  |  |
| No | 8192 | (55.4) |  |
| Yes | 6596 | (44.6) |  |
| Psychiatric disorder<sup>4</sup> |  |  |  |
| No | 10680 | (74.0) |  |
| Yes | 3747 | (26.0) |  |
| Essential service worker |  |  |  |
| No | 12192 | (84.1) |  |
| Yes | 2307 | (15.9) |  |
| Healthcare worker |  |  |  |
| No | 13867 | (95.6) |  |
| Yes | 632 | (4.4) |  |
| Parent of children <18 years |  |  |  |
| No | 11490 | (78.5) |  |
| Yes | 3145 | (21.5) |  |
| Results of COVID-19 test |  |  |  |
| Others | 14702 | (99.0) |  |
| COVID-19 positive | 144 | (1.0) |  |
| History of getting flu vaccine |  |  |  |
| < 3 times in past 5 years | 8348 | (57.0) |  |
| ≥ 3-5 times in past 5 years | 6304 | (43.0) |  |

1. Western provinces: British Columbia, Alberta, Saskatchewan, and Manitoba 2. Atlantic provinces: Nova Scotia, New Brunswick, Prince Edward Island, and Newfoundland/Labrador 3. High-risk health conditions: cardiovascular disease, chronic respiratory disease, diabetes, obesity, cancer, autoimmune disease 4. Psychiatric disorders: any mood and/or anxiety disorder and dementia

### Estimates of vaccine hesitancy and changes over time

Rates of vaccine hesitancy across time/survey round are presented in **Figure 1**. Overall, 42.2% of respondents reported vaccine hesitancy over the course of the study period, though we observed significant variations in vaccine hesitancy rates over time (survey 1: 36.8%, survey 2: 44.6%; survey 3: 52.9%, survey 4: 39.6%, survey 5: 36.9%). As can be seen in Figure 1, vaccine hesitancy was lowest during surveys 1 (April 2020) and 5 (March 2021), and highest during survey 3 (November 2020).

**Figure 1.**
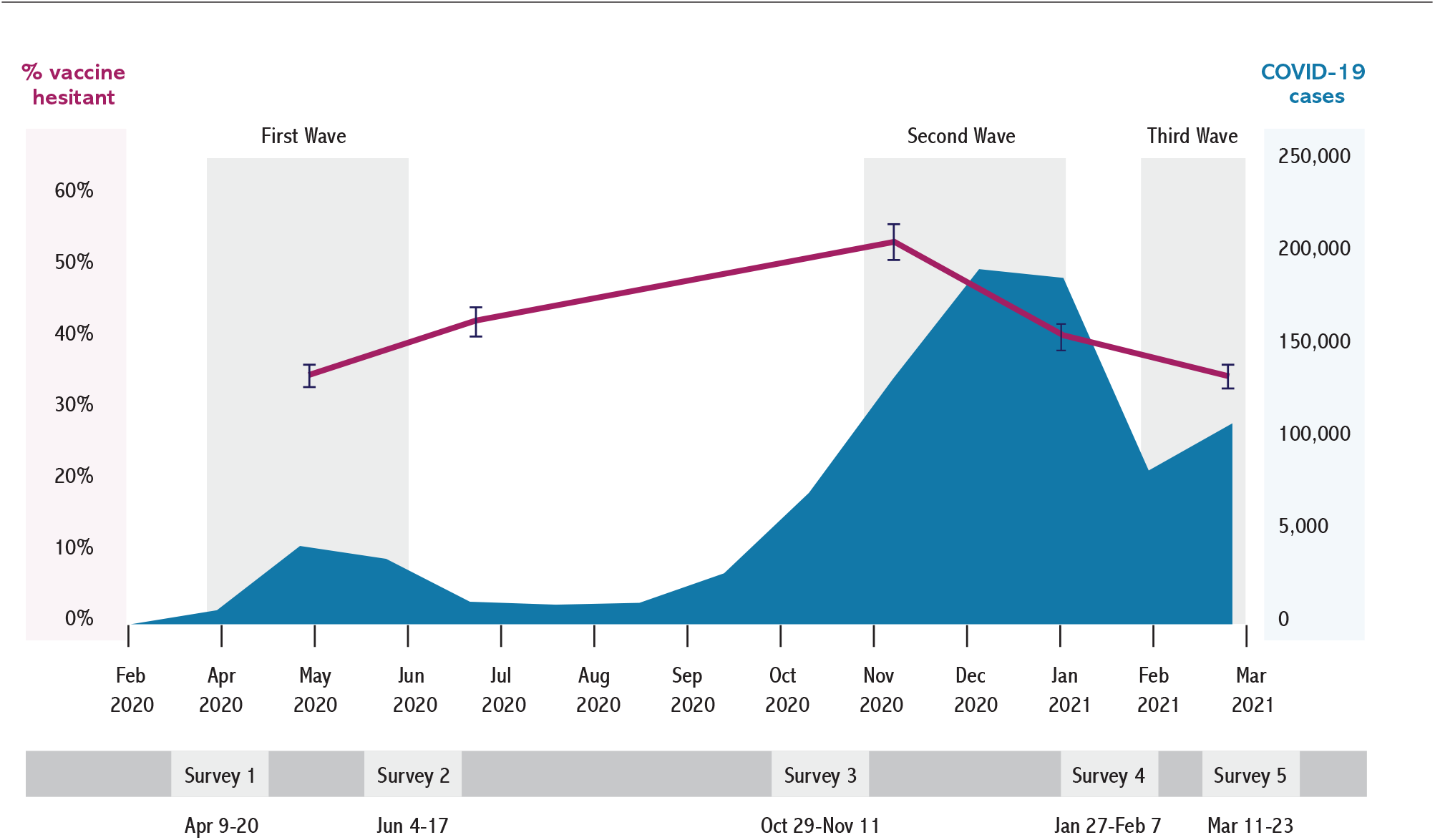
Rates of vaccine hesitancy across the five surveys/time points.

Participant characteristics presented as a function of vaccine hesitancy status across all surveys/time points are presented in **Figure 2** (individual survey data can be found in **Supplement Table S2**). Across all surveys, rates of vaccines hesitancy were significantly higher among younger age groups (< 25 years and 26-50 years compared to those aged 50+), non-Whites, those currently employed, those reporting less than $60,000 in annual family income, and those living in Western provinces (British Columbia, Alberta, Saskatchewan and Manitoba) and Ontario compared to Quebec and the Atlantic provinces. In addition, rates of vaccine hesitancy were significantly higher among those without a health risk condition, those identifying as essential workers, those identifying as healthcare workers, and parents of children under 18. Finally, rates of vaccine hesitancy were significantly higher among those reporting getting the flu vaccine less than three times in the past five years (all p’s <0.05).

**Figure 2.**
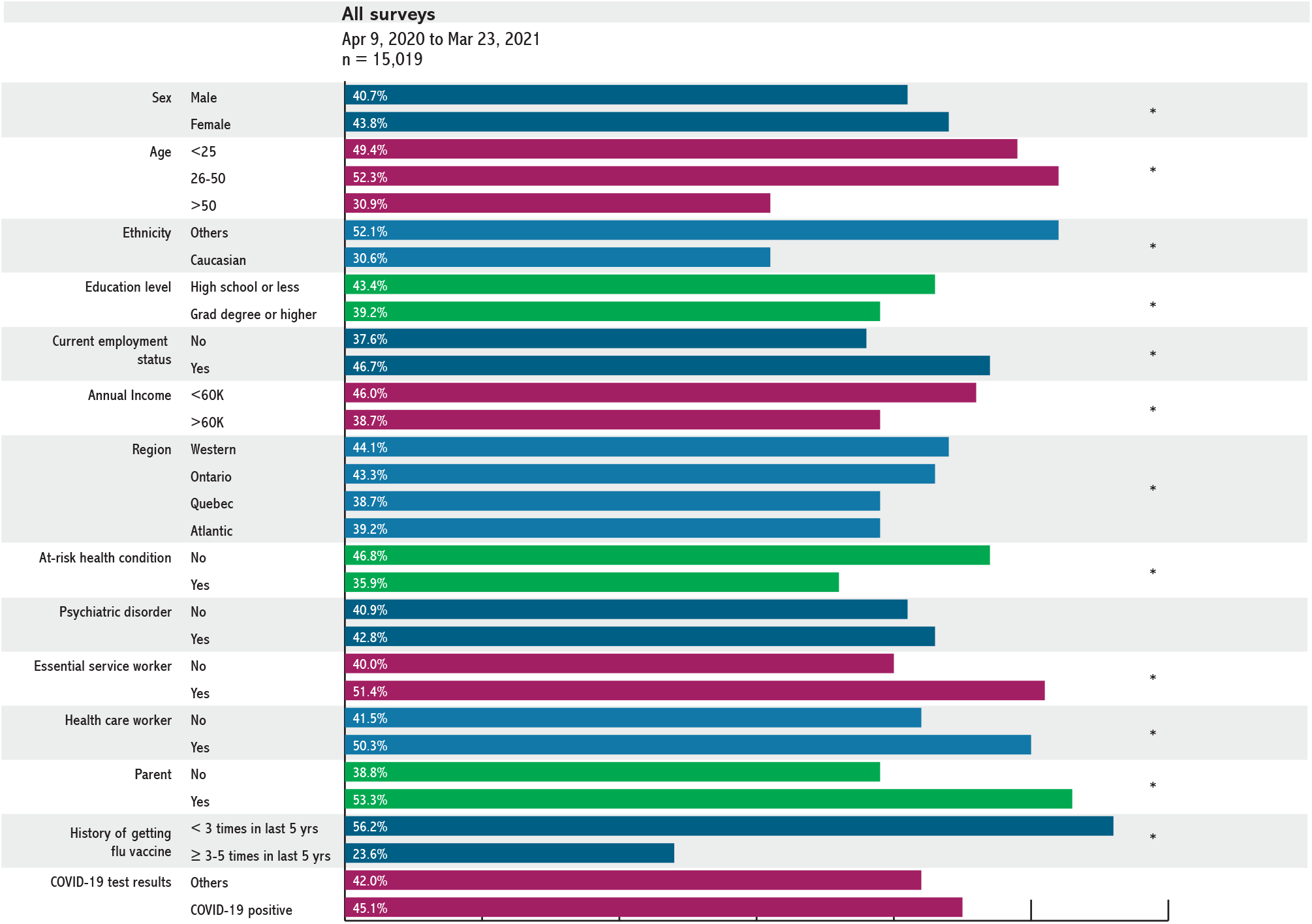
Participant characteristics presented as a function of being hesitant* vs. extremely likely to get a COVID-19 vaccine across the three surveys: univariate analyses. **\* Hesitant:** those reporting being “somewhat likely”, “unlikely” or “extremely unlikely” to seek out the COVID-19 vaccine **Western provinces:** British Columbia, Alberta, Saskatchewan, and Manitoba **Atlantic provinces:** Nova Scotia, New Brunswick, Prince Edward Island, and Newfoundland/Labrador **High-risk health conditions:** cardiovascular disease, chronic respiratory disease, diabetes, obesity, cancer, autoimmune disease **Psychiatric disorders:** any mood and/or anxiety disorder and dementia

### Sociodemographic predictors of vaccine hesitancy

Multivariable logistic regression analyses examining associations between vaccine hesitancy and sociodemographic and clinical variables across all surveys/time points are presented in **Table 2**. Partially adjusted analyses revealed that women were 19% more likely to be vaccine hesitant (OR_padj_ 1.19, 95% CI 1.08 – 1.32), those aged less than 25 years (OR_padj_ 2.07, 95% CI 1.74 – 2.46) and 26-50 years (OR_padj_ 2.,41 95% CI 2.16 – 2.69) were 2.07 times and 2.41 times more likely to be hesitant compared to those aged 51 and over, and those who identified as non-White were 1.3 times more likely to be vaccine hesitant compared to Whites (OR_adj_ 1.30, 95% CI 1.14 – 1.49). Fully adjusted analyses revealed that in addition to women, younger age groups, and non-Whites, those with high school or less education were 1.15 times more likely to be vaccine hesitant compared to those with graduate or post-graduate degrees (OR_adj_ 1.15, 95% CI 1.041- 1.28), those earning less than $60,000 per year in household income were 1.42 times more likely to be vaccine hesitant that those earning $60,000 or more (OR_adj_ 1.42, 95% CI 1.26-1.61), essential and healthcare workers were 1.44 (OR_adj_ 1.44, 95% CI 1.21 – 1.71) and 1.35 (OR_adj_ 1.35, 95% CI 1.04 – 1.75) times more likely to be vaccine hesitant respectively, compared to those not in those fields. Finally, parents of children under 18 were 1.51 times more likely to be vaccine hesitant compared to non-parents (OR_adj_ 1.51, 95% CI 1.30 – 1.75), and those reporting getting the flu vaccine three times or more in the past five years were 73% less likely to be vaccine hesitant compared to those reporting getting the flu vaccine less than three times in the past five years (OR_adj_ 0.27, 95% CI 0.23 – 0.30).

**Table 2.**
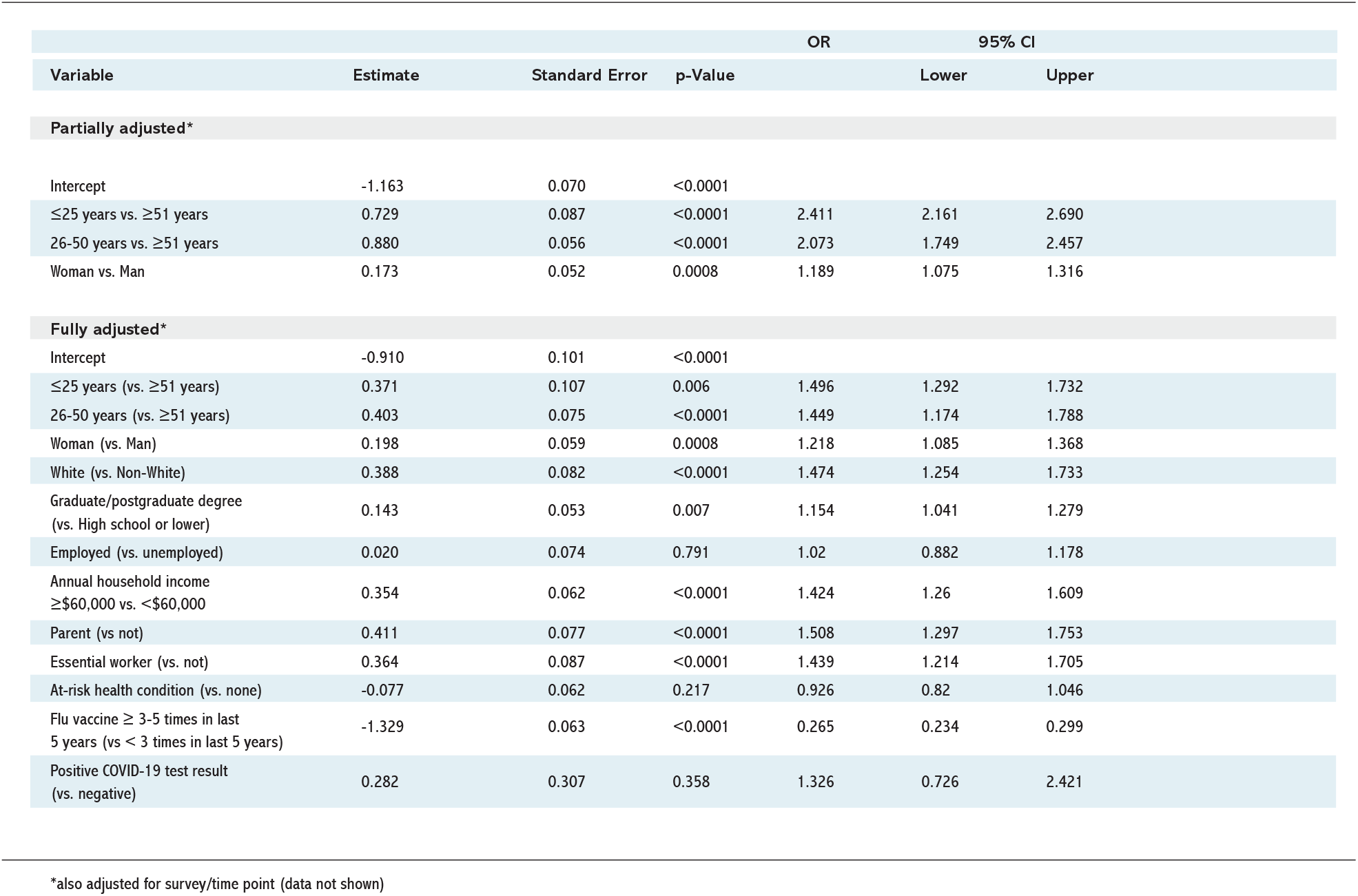
Multivariate associations between sociodemographic characteristics and COVID-19 vaccine hesitancy.

|  |  |  |  | OR | 95% CI |  |
| --- | --- | --- | --- | --- | --- | --- |
| Variable | Estimate | Standard Error | p-Value |  | Lower | Upper |
| Partially adjusted* |  |  |  |  |  |  |
| Intercept | -1.163 | 0.070 | <0.0001 |  |  |  |
| ≤25 years vs. ≥51 years | 0.729 | 0.087 | <0.0001 | 2.411 | 2.161 | 2.690 |
| 26-50 years vs. ≥51 years | 0.880 | 0.056 | <0.0001 | 2.073 | 1.749 | 2.457 |
| Woman vs. Man | 0.173 | 0.052 | 0.0008 | 1.189 | 1.075 | 1.316 |
| Fully adjusted* |  |  |  |  |  |  |
| Intercept | -0.910 | 0.101 | <0.0001 |  |  |  |
| ≤25 years (vs. ≥51 years) | 0.371 | 0.107 | 0.006 | 1.496 | 1.292 | 1.732 |
| 26-50 years (vs. ≥51 years) | 0.403 | 0.075 | <0.0001 | 1.449 | 1.174 | 1.788 |
| Woman (vs. Man) | 0.198 | 0.059 | 0.0008 | 1.218 | 1.085 | 1.368 |
| White (vs. Non-White) | 0.388 | 0.082 | <0.0001 | 1.474 | 1.254 | 1.733 |
| Graduate/postgraduate degree<br>(vs. High school or lower) | 0.143 | 0.053 | 0.007 | 1.154 | 1.041 | 1.279 |
| Employed (vs. unemployed) | 0.020 | 0.074 | 0.791 | 1.02 | 0.882 | 1.178 |
| Annual household income<br>≥\$60,000 vs. <\$60,000 | 0.354 | 0.062 | <0.0001 | 1.424 | 1.26 | 1.609 |
| Parent (vs not) | 0.411 | 0.077 | <0.0001 | 1.508 | 1.297 | 1.753 |
| Essential worker (vs. not) | 0.364 | 0.087 | <0.0001 | 1.439 | 1.214 | 1.705 |
| At-risk health condition (vs. none) | -0.077 | 0.062 | 0.217 | 0.926 | 0.82 | 1.046 |
| Flu vaccine ≥ 3-5 times in last<br>5 years (vs < 3 times in last 5 years) | -1.329 | 0.063 | <0.0001 | 0.265 | 0.234 | 0.299 |
| Positive COVID-19 test result<br>(vs. negative) | 0.282 | 0.307 | 0.358 | 1.326 | 0.726 | 2.421 |

\*also adjusted for survey/time point (data not shown)

### Psychological predictors of vaccine hesitancy

Perceptions of the importance of engaging in infection prevention behaviours preventive across the five surveys/time points is presented in **Figure 3**. Overall, 76% of respondents reported believing that engaging in infection prevention behaviours was extremely important, though we observed significant variations in perceived importance over time. Perceived importance was highest at survey 1 (87%), which then dropped to 71.3% by survey 2 and remained generally stable across survey 3 (74.5%), 4 (75.7%) and survey 5 (71.3%). Concern trends generally followed a similar pattern: mean values for each concern type were highest at survey 1, and dropped significantly by survey 2 and remained generally stable across survey 3 to 5 (p<.0001 for trend, see **Figure 3**). Across all five surveys/time points, respondents reported having the greatest concerns about the social and economic impacts of the pandemic (M= 3.18, SD 0.76), followed by health concerns (M=2.98, SD=0.86) and personal financial concerns (M=2.43, SD=1.08).

**Figure 3.**
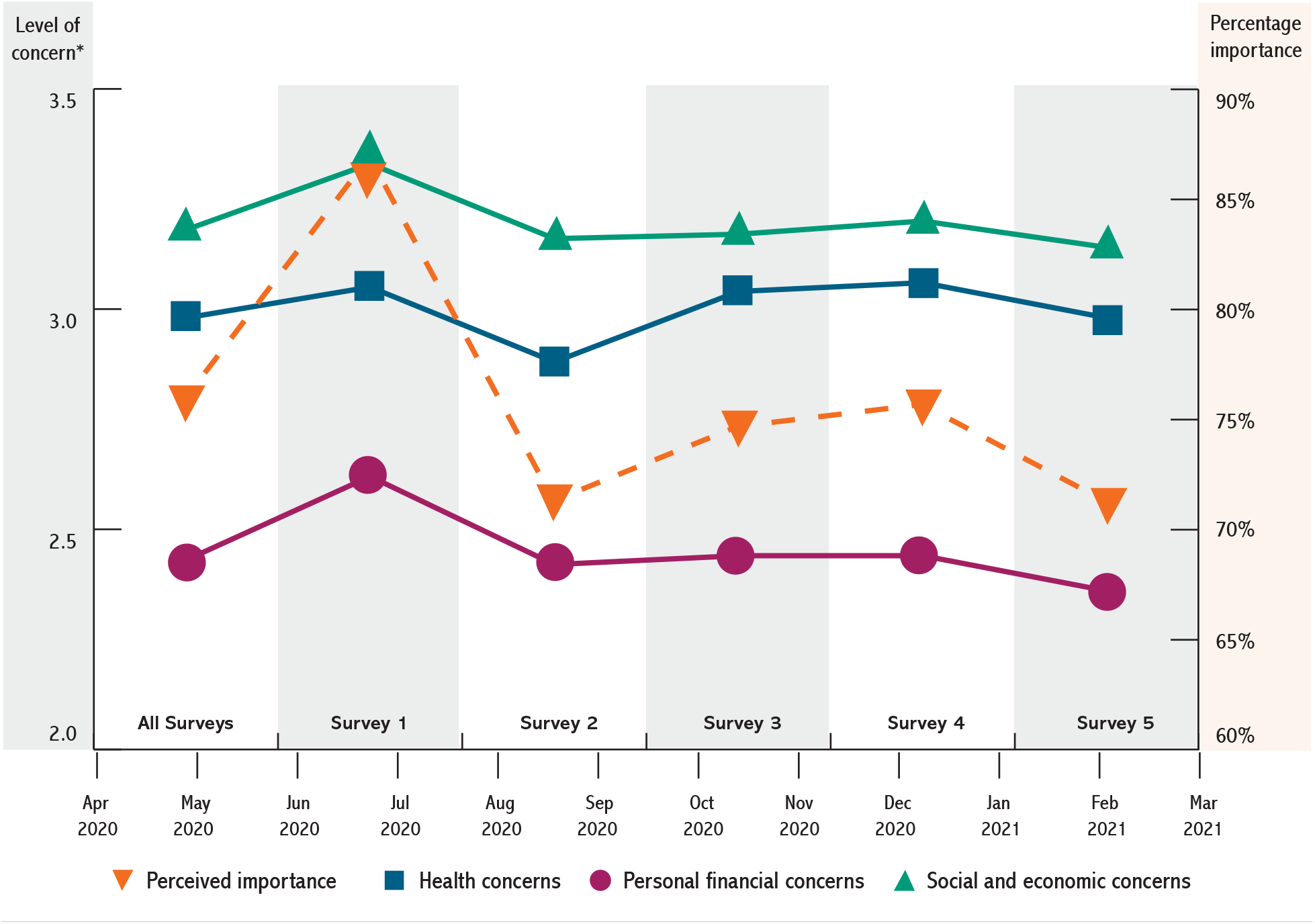
Perceptions of the importance of engaging in infection prevention behaviours (percentage of respondents reporting “extremely important”, dashed line) and mean COVID-19 concern levels (solid lines) across the five surveys/time points. * Mean concern level (scale: 0-4)

Partially and fully adjusted multivariate logistic regression analyses examining associations between vaccine hesitancy and perceived importance of engaging in infection prevention behaviours and COVID-19- related concern types across all surveys/time points are presented in **Tables 3 and 4**. Respondents who perceived engaging in infection prevention behaviours to be extremely important were 78% (partially adjusted) and 77% (fully adjusted) less likely to be vaccine hesitant than those who believed engaging in these behaviours was only somewhat, not very, or not at all important (OR_padj_ 0.22, 95% CI 0.19 – 0.25 and OR_adj_ 0.23, 95% CI 0.20 – 0.27, respectively). Although social and economy concerns were the most endorsed by respondents, they were not predictive of vaccine hesitancy in partially or fully adjusted analyses. However, health concerns were associated with a 58% (partially adjusted) and 54% (fully adjusted) reduced odds of vaccine hesitancy (OR_padj_ 0.42, 95% CI 0.39 – 0.46 and OR_adj_ 0.46, 95% CI 0.42 – 0.50, respectively), while having high personal financial concerns was associated with a 1.41 and 1.33 times greater odds of vaccine hesitancy in partially (OR_padj_ 1.41, 95% CI 1.32 – 1.49) and fully adjusted (OR_adj_ 1.33, 95% CI 1.25-1.43) models.

**Table 3.**
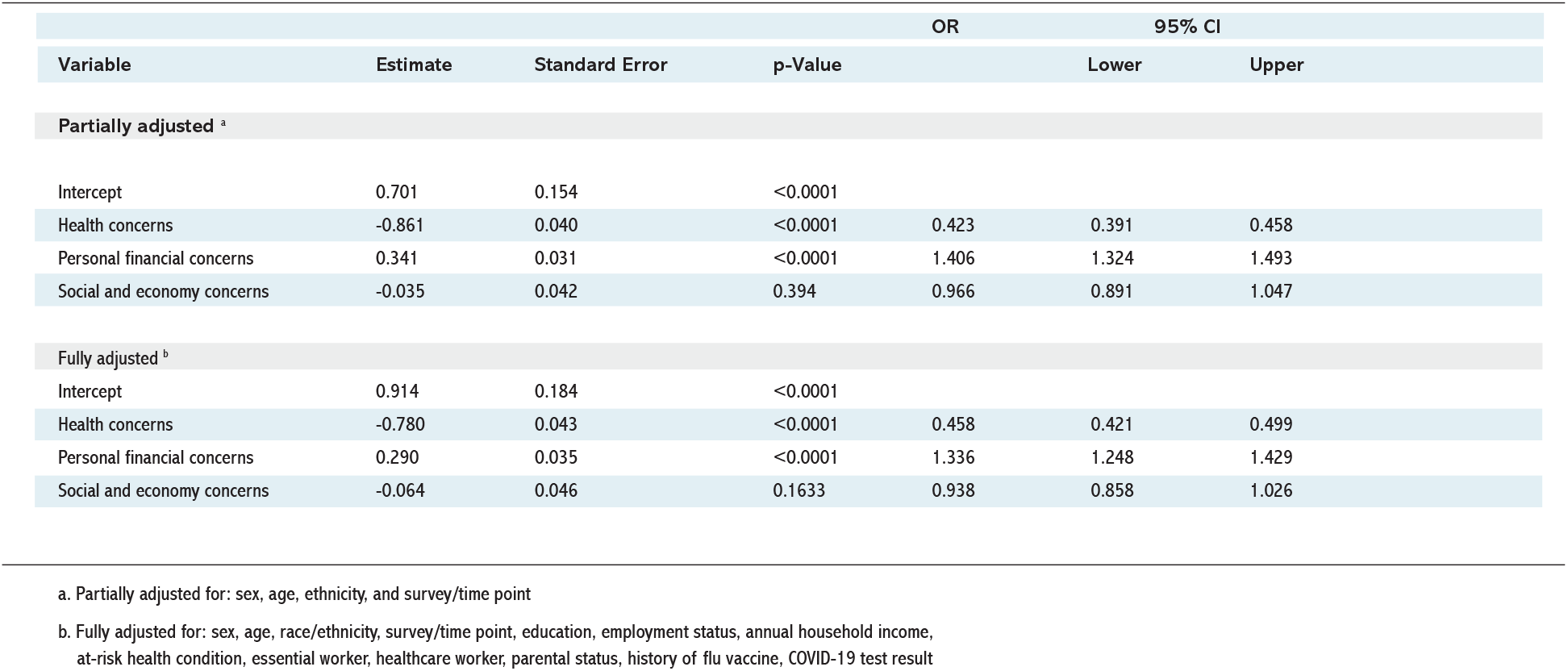
Multivariate logistic regression model estimating the association between COVID-19-related concerns and vaccine hesitancy.

**Table 4.** Multivariate logistic regression model estimating the association between perceived importance of COVID-19 prevention measures and vaccine hesitancy.

|  |  |  |  | OR | 95% CI |  |
| --- | --- | --- | --- | --- | --- | --- |
| Variable | Estimate | Standard Error | p-Value |  | Lower | Upper |
| **Partially adjusted <sup>a</sup>** |  |  |  |  |  |  |
| Intercept | 0.168 | 0.092 | 0.0665 |  |  |  |
| Perceived importance: |  |  |  |  |  |  |
| Very important vs. others ¶ | -1.536 | 0.065 | <0.0001 | 0.215 | 0.19 | 0.245 |
| **Fully adjusted <sup>b</sup>** |  |  |  |  |  |  |
| Intercept | 0.371 | 0.123 | 0.0026 |  |  |  |
| Perceived importance: |  |  |  |  |  |  |
| Very important vs. others ¶ | -1.462 | 0.073 | <0.0001 | 0.232 | 0.201 | 0.267 |

a. Partially adjusted for: sex, age, race/ethnicity, and survey/time point
b. Fully adjusted for: sex, age, ethnicity, survey/time point, education, employment status, annual household income, at-risk health condition, essential worker, healthcare worker, parental status, history of flu vaccine, COVID-19 test result
¶. Others: Somewhat important, unimportant or not at all important

## Discussion

The present study analyzed Canadian survey data from five age, sex and province-weighted population-based samples to describe vaccine intentions between April 2020 and March 2021 and their correlates. Over 40% of Canadians reported some degree of vaccine hesitancy over the course of the study period. Vaccine hesitancy was lowest during pandemic waves one and three, and highest during pandemic wave 2, just prior to vaccine approval in Canada (December 2020). These results are consistent with data from the US covering the same time period, which also demonstrated significant increases in vaccine hesitancy between April and December 2020 among 8,167 online respondents in the Understanding America Study.^6^ These results are also aligned with those of a study conducted by the World Economic Forum, which reported a decline in positive vaccine intentions between August (77%) and October 2020 (73%) among 18,526 respondents from 15 countries (including 1,000 from Canada)^17^.

### Profile of Canadians who are vaccine hesitant

We examined the profile of Canadians who were more likely to report being vaccine hesitant, and found that in fully adjusted analyses (including survey/time point), women, younger individuals (aged 50 and younger), non-Whites, those with lower levels of education (high school or less), and those reporting lower annual household incomes (less than $60,000/year) were significantly more likely to report being vaccine hesitant over the study period. Overall, this profile is consistent with the results of similar studies in Canada and other Western nations (e.g., USA, UK, France, Italy, Germany, and Australia)^18-28^ suggesting a robust phenomenon of higher vaccine hesitancy among women, younger individuals, non-Whites, and those of lower socioeconomic status.

The reasons for the lower vaccine intentions among women remains poorly understood, and seems paradoxical given evidence that women are more adherent to COVID-19 prevention measures in general.^15,29^ Some speculate it might be related to their tendency to have greater health risk perceptions in general,^30^ which may lead to heightened fears of experiencing vaccine side effects compared to men, resulting in less willingness to get vaccinated. These fears may not be completely unfounded, in light of evidence showing that women tend to have stronger immune reactions to vaccines than men, which may lead to more adverse events following vaccination.^31,32^ More recent data suggests that women may be more reluctant to get vaccinated due to reproductive factors, as women who are pregnant or planning to get pregnant appear to be delaying vaccination due to safety concerns affecting the fetus.^33,34^ Given evidence to suggest that pregnancy in the presence of COVID-19 may confer increased risk for severe illness, hospitalization and intensive care unit admission, and preliminary findings of no obvious safety concerns among pregnant women who received mRNA vaccines^35^, addressing vaccine hesitancy in this group will be important for protecting this vulnerable population.

Contrary to women, younger adults may be less willing to get vaccinated due to *lower* COVID-19 risk perception compared to older adults.^36^ These perceptions may have been fueled by early reports of lower risks of COVID-19 hospitalization and complications among younger age groups.^37^ While overall COVID-19-related mortality among those under age 20 remains low (proportion of all-cause deaths attributed to COVID-19 has been estimated to be 0.48%^38^), those aged 20 to 59 have accounted for 63% of all infections and 30% of all hospitalizations in Canada since the start of the pandemic.^39^ This suggests that this age group remains an important vector community virus transmission, and a need to optimize vaccination uptake in this age cohort.

Our results also revealed lower vaccine intentions among non-Whites, those with high school or less education, and those with annual household incomes less than $60,000CAD/year (below the poverty line in Canada).^40^ These results are consistent with those from previous studies in the US^18,20,21,41^, Australia^25^, and across Europe^26,27,42,43^. Results of greater vaccine hesitancy among people of color is a cause for concern, given that these individuals are more likely to work in industries worst affected by the COVID-19 pandemic, such as food and beverage, hospitality, and long-term care services.^44^ Reasons for higher rates of hesitancy among these groups may include lower health literacy^45^ and lack of trust in vaccines and the healthcare system,^46^ the latter of which may be exacerbated by low representation of people of color in vaccine trials and experience with discrimination and systemic racism.^47^ Clearly, greater efforts need to be made to motivate and enable those from racial and ethnic minority groups to get vaccinated.

We also identified two important groups of individuals at greater risk of being vaccine hesitant: essential and healthcare workers. Evidence of greater hesitancy among essential and healthcare workers was both surprising and a cause of concern, given that they are the individuals most likely to be exposed, and expose others to COVID-19. However, our results do seem to be in line with US data from a survey of 16,970 employed adults in the US showing that those working in essential service sectors (i.e., leisure and hospitality, manufacturing, construction, retail, transportation, and food and beverage) had the highest rates of vaccine hesitancy (45% to 54%) compared to non-essential sectors like technology (25%), financial services (26%), public administration (36%) and entertainment (37%).^48^ Our findings of high vaccine hesitancy among healthcare workers is also consistent with other studies both within^49^ and outside^50-52^ of Canada. Though we were not able to determine what types of healthcare workers are more likely to be vaccine hesitant, data from previous reports suggests this is more common among female healthcare workers,^49,50,53^ as well as nurses and paramedical professionals rather than physicians or health administrators.^51-53^ While the reasons for vaccine hesitancy among healthcare workers remain poorly understood, available evidence suggests their hesitancy is linked to vaccine novelty and concerns about safety.^49,52^ Further research is needed to identify barriers to vaccination among essential and healthcare workers due to their high risk of virus exposure and transmission.

There were two additional findings from our analyses that warrant discussion. The first is that vaccine hesitancy was higher among those with an inconsistent history of getting the flu vaccine. This is consistent with previous reports^19,41,43,54,55^, and suggests that having favorable vaccine attitudes and behaviours in general is associated with greater likelihood of getting vaccinated against COVID-19. The other finding is that parents of children under age 18 were 1.5 times more likely to be vaccine hesitant compared to non-parents. Given the impending approval of vaccines among 5-11 year-olds in Canada, this finding is a cause for concern, and consistent with at least one study out of the UK that also found that parents of young children were more likely to report vaccine hesitancy or refusal.^56^ The reasons for this are remain poorly understood, but may reflect more general trends of parental hesitancy to vaccinate their children against common infectious diseases (e.g., mumps, measles, pertussis).^57^ Given that COVID-19 infection rates are currently highest among school-aged children in Canada,^58^ parents represent an important target for vaccination. Further research is needed to understand the reasons for vaccine hesitancy in this group and the impact of personal vaccine hesitancy on their willingness to get their children vaccinated against COVID-19, in order to optimise vaccination rates in this vulnerable group.

### Psychological predictors of vaccine hesitancy

In addition to sociodemographic predictors, we also assessed psychological predictors of vaccine hesitancy. One of the strongest predictors of positive vaccine intentions was the extent to which Canadians believed engaging in preventive health behaviours (e.g., vaccination) was important for reducing virus transmission. Those who believed that engaging in preventive health behaviours (like getting vaccinated) was ‘extremely important’ were 77% less likely to be vaccine hesitant after adjustment for covariates including sociodemographics and survey period/time point. This finding is consistent with previous reports linking high perceived benefits (of getting vaccinated) to positive vaccine intentions,^59^ highlighting the need for vaccination campaigns to clearly and consistently emphasize how the benefits of getting vaccinated far outweigh any risks. We also found that different types of COVID-19-related concerns were important determinants of vaccine hesitancy. Interestingly, even though social and economy concerns were the most highly endorsed at each survey/time point, only high health-related concerns and personal financial concerns were significant predictors of vaccine hesitancy – but not in the same direction. In fact, we found that those with high health concerns (i.e., concerned about becoming infected and/or infecting others) were 54% less likely to be vaccine hesitant, while those with high concerns about their personal financial situation (e.g., were worried about job and income loss or not having enough money to feed their family) were 1.33 times *more likely* to report being vaccine hesitant. Results linking high health concerns to lower vaccine hesitancy are consistent with those of other studies in Canada, the US, Australia, and Europe^18,20,21,23,26,55,60-63^, and provide further support of the need for vaccination campaigns to highlight how getting vaccinated is going to be health protective. However, to our knowledge, this is the first study to date to observe a link between high personal financial concerns and increased vaccine hesitancy, and suggests that those whose livelihoods were negatively impacted by the virus may be less willing or able to get vaccinated. Further research is needed to determine the extent to which this reflects a lack of motivation or desire to get vaccinated, or a perceived inability to get vaccinated due to practical barriers or limitations (e.g., lack of access to paid leave to get vaccinated).

### Limitations and strengths

This study should be interpreted in light of some methodological limitations. First, although we included large, national samples of Canadians with representation across age, sex, and province, the absolute number of participants in certain provinces (e.g., Atlantic) was lower, making inter-provincial comparisons difficult. Second, the survey was only available in English and French, which may have led to an underrepresentation of certain non-native English or French speaking groups. Further, our surveys included fewer people of color, which may reflect participation on online panels, so results might not generalize as well to non-Whites. Third, since the surveys were voluntary and participants were drawn from a polling firm’s subject pool, participation may have been subject to some degree of selection bias. Fourth, though this study presents data depicting vaccine intentions over time, it was drawn from three separate cohorts of online panels, so data reflect trends in vaccine intentions over time but not in the same individuals. Finally, data were self-reported, which may have been subject to social desirability bias.^64^ However, the fact that the surveys were anonymous likely mitigated this limitation.

Despite some limitations, this study also had a number of important strengths. The study included a large sample size, respondents were well distributed across provincial regions, age groups, employment status and income compared to census data available through Statistics Canada, and there were equal proportions of men and women. This study also collected data during peak lockdown of the first wave (April 2020) through to end the third wave (end of March 2021) when vaccines started becoming available in Canada. This allowed for the assessment of changes in vaccine intentions over time across three critical waves of the pandemic in Canada. We used conducted principal component analysis to determine the structure of our concerns module, which was found to have excellent internal consistency, which is important for ensuring the validity of our results linking concern types to vaccine hesitancy. Finally, results reflect a sub-analysis of Canadian representative data from the iCARE study, which has collected data from more than 100,000 people from 190 countries to date alongside ongoing efforts to collect similarly representative samples in eight other countries (see: www.icarestudy.com). This will facilitate comparisons with international datasets to contribute important evidence to support the development and implementation of COVID-19 vaccine policy strategies worldwide.

## Conclusions

Over 40% of Canadians reported some degree of vaccine hesitancy between April 2020 and March 2021. Vaccine hesitancy was lowest during pandemic waves one and three, and highest during pandemic wave 2, just prior to vaccine approval in Canada. Women, individuals aged 50 and younger, non-Whites, those with high school education or less, and those with annual household incomes below the poverty line in Canada (i.e., $60,000) were significantly more likely to report being vaccine hesitant over the study period. Three important groups of Canadians were identified as being vaccine hesitant: essential and healthcare workers, parents of children under the age of 18, and those without a previous history of flu vaccination. Finally, perceived importance of engaging in infection prevention behaviours (like vaccination) and having high COVID-19-related health concerns were predictive of lower levels of vaccine hesitancy, whereas having high COVID-19-related personal financial concerns was predictive of higher levels of vaccine hesitancy. Overall, results point to the importance of targeting vaccine efforts to subgroups who may be socioeconomically disadvantaged, who also happen to be disproportionately represented in essential service occupations including healthcare. Finally, vaccine messaging should emphasize how the benefits of getting vaccinated (particularly to health) far outweigh the risks, particularly those associated with personal financial losses. Future research is needed to monitor ongoing changes in vaccine intentions and behaviour, as well as to better understand motivators and facilitators of vaccine acceptance, particularly among vulnerable groups.

## Data Availability

All data produced in the present study are available upon reasonable request to the authors.

## Acknowledgements

The primary source of funding for the iCARE study has been primarily through re-directed funding associated with Montreal Behavioural Medicine Centre, including funds from the Canada Research Chairs Program (950- 232522, Chair holder: Dr. Kim L. Lavoie), a Canadian Institutes of Health Research-Strategy for Patient Oriented Research Mentoring Chair (SMC-151518, PI: Dr. Simon L. Bacon), a Fonds de Recherche du Québec: Santé Chair (251618, PI: Dr. Simon L. Bacon), and a Fonds de Recherche du Québec: Santé Senior Research Award (34757, PI: Dr. Kim L Lavoie). Addition support has been provided by the Canadian Institutes of Health Research (MS3- 173099, co-PI’s: Simon L. Bacon & Kim L. Lavoie) and the Fonds de Recherche du Québec: Société et Culture (2019-SE1-252541, PI: Dr. Simon L. Bacon). None of the funders were involved in the study design. Dr. Lavoie is a member of the Canadian COVID-19 Expert Advisory Panel (Health Canada). Dr. Presseau is a member of the Ontario Immunization Advisory Committee (Public Health Ontario) and a member of the Ontario COVID-19 Science Advisory Table. We also acknowledge the support from our MBMC iCARE Team, particularly administrative support by Mr. Guillaume Lacoste and Genevieve Szczepanik, and analytic support by Ms. Mariam Atoui and Julian Esse Atto.

## iCARE Study Collaborators

**Lead investigators:** Kim L. Lavoie, PhD, University of Quebec at Montreal (UQAM) and CIUSSS-NIM, CANADA; Simon L. Bacon, PhD, Concordia University and CIUSSS-NIM, CANADA. **Collaborators** (in alphabetical order by country): ABU DHABI: Zahir Vally, PhD, United Arab Emirates University; ARGENTINA: Analía Verónica Losada, PhD, University of Flores; AUSTRALIA: Jacqueline Boyle, PhD, Monash University; Joanne Enticott, PhD, Monash University; Shajedur Rahman Shawon, PhD, Centre for Big Data Research in Health, UNSW Medicine; Helena Teede, MD, Monash University; AUSTRIA: Alexandra Kautzky-Willer, MD, Medizinische Universität Wien; BANGLADESH: Arobindu Dash, MS, International University of Business, Agriculture & Technology; BRAZIL: Marilia Estevam Cornelio, PhD, University of Campinas; Marlus Karsten, Universidade do Estado de Santa Catarina - UDESC; Darlan Lauricio Matte, PhD, Universidade do Estado de Santa Catarina - UDESC; CANADA: Ahmed Abou-Setta, PhD, University of Manitoba; Shawn Aaron, PhD, Ottawa Hospital Research Institute; Angela Alberga, PhD, Concordia University; Tracie Barnett, PhD, McGill University; Silvana Barone, MD, Université de Montréal; Ariane Bélanger-Gravel, PhD, Université Laval; Sarah Bernard, PhD, Université Laval; Lisa Maureen Birch, PhD, Université Laval; Susan Bondy, PhD, University of Toronto - Dalla Lana School of Public Health; Linda Booij, PhD, Concordia University; Roxane Borgès Da Silva, PhD, Université de Montréal; Jean Bourbeau, MD, McGill University; Rachel Burns, PhD, Carleton University; Tavis Campbell, PhD, University of Calgary; Linda Carlson, PhD, University of Calgary; Kim Corace, PhD, University of Ottawa; Olivier Drouin, MD, CHU Sainte-Justine/Université de Montréal; Francine Ducharme, MD, Université de Montréal; Mohsen Farhadloo, Concordia University; Carl Falk, PhD, McGill University; Richard Fleet MD, PhD, Université Laval; Michel Fournier, MSc, Direction de la Santé Publique de Montréal; Gary Garber, MD, University of Ottawa/Public Health Ontario; Lise Gauvin, PhD, Université de Montréal; Jennifer Gordon, PhD, University of Regina; Roland Grad, MD, McGill University; Samir Gupta, MD, University of Toronto; Kim Hellemans, PhD, Carleton University; Catherine Herba PhD, UQAM; Heungsun Hwang, PhD, McGill University; Jack Jedwab, PhD, Canadian Institute for Identities and Migration and the Association for Canadian Studies; Keven Joyal-Desmarais, PhD, Concordia University; Lisa Kakinami, PhD, Concordia University; Eric Kennedy, PhD, York University; Sunmee Kim, PhD, University of Manitoba; Joanne Liu, PhD, McGill University; Colleen Norris, PhD, University of Alberta; Sandra Pelaez, PhD, Université de Montréal; Louise Pilote, MD, McGill University; Paul Poirier, MD, Université Laval; Justin Presseau, PhD, University of Ottawa; Eli Puterman, PhD, University of British Columbia; Joshua Rash, PhD, Memorial University; Paula AB Ribeiro, PhD, MBMC; Mohsen Sadatsafavi, PhD, University of British Columbia; Paramita Saha Chaudhuri, PhD, McGill University; Jovana Stojanovic, PhD, Concordia University; Eva Suarthana, MD, PhD, Université de Montréal/McGill University; Sze Man Tse, MD, CHU Sainte-Justine; Michael Vallis, PhD, Dalhousie University; CHILE: Nicolás Bronfman Caceres, PhD, Universidad Andrés Bello; Manuel Ortiz, PhD, Universidad de La Frontera; Paula Beatriz Repetto, PhD, Universidad Católica de Chile; COLOMBIA: Mariantonia Lemos-Hoyos, PhD, Universidad EAFIT; CYPRUS: Angelos Kassianos, PhD, University of Cyprus; DENMARK: Naja Hulvej Rod, PhD, University of Copenhagen; FRANCE: Mathieu Beraneck, PhD, Université de Paris; CNRS; Gregory Ninot, PhD, Université de Montpellier; GERMANY: Beate Ditzen, PhD, Heidelberg University; Thomas Kubiak, PhD, Mainz University; GHANA: Sam Codjoe MPhil,MSc, University of Ghana; Lily Kpobi, PhD, University of Ghana; Amos Laar, PhD, University of Ghana; INDIA: Naorem Kiranmala Devi, PhD, University of Delhi; Sanjenbam Meitei, PhD, Manipur University; Suzanne Tanya Nethan, MDS, ICMR-National Institute of Cancer Prevention & Research; Lancelot Pinto, MD, PhD, Hinduja Hospital and Medical Research Centre; Kallur Nava Saraswathy, PhD, University of Delhi; Dheeraj Tumu, MD, World Health Organization (WHO); INDONESIA: Silviana Lestari, MD, PhD, Universitas Indonesia; Grace Wangge, MD, PhD, SEAMEO Regional Center for Food and Nutrition; IRELAND: Molly Byrne, PhD, National University of Ireland, Galway; Hannah Durand, PhD, National University of Ireland, Galway; Jennifer McSharry, PhD, National University of Ireland, Galway; Oonagh Meade, PhD, National University of Ireland, Galway; Gerry Molloy, PhD, National University of Ireland, Galway; Chris Noone, PhD, National University of Ireland, Galway; ISRAEL: Hagai Levine, MD, Hebrew University; Anat Zaidman-Zait, PhD, Tel-Aviv University; ITALY: Stefania Boccia, PhD, Università Cattolica del Sacro Cuore; Ilda Hoxhaj, MD, Università Cattolica del Sacro Cuore, Stefania Paduano, MSc, PhD, University of Modena and Reggio Emilia; Valeria Raparelli, PhD, Sapienza - University of Rome; Drieda Zaçe, MD, MSc, PhDc, Università Cattolica del Sacro Cuore; JORDAN: Ala’S Aburub, PhD, Isra University; KENYA: Daniel Akunga, PhD, Kenyatta University; Richard Ayah, PhD, University of Nairobi, School Public Health; Chris Barasa, MPH, University of Nairobi, School Public Health; Pamela Miloya Godia, PhD, University of Nairobi; Elizabeth W. Kimani-Murage, PhD, African Population and Health Research Center; Nicholas Mutuku, PhD, University of Kenya; Teresa Mwoma, PhD, Kenyatta University; Violet Naanyu, PhD, Moi University; Jackim Nyamari, PhD, Kenyatta University; Hildah Oburu, PhD, Kenyatta University; Joyce Olenja, PhD, University of Nairobi; Dismas Ongore, PhD, University of Nairobi; Abdhalah Ziraba, PhD, African Population and Health Research Center; MALAWI: Chiwoza Bandawe, PhD, University of Malawi; MALAYSIA: Loh Siew Yim, PhD, Faculty of medicine, University of Malaya; NEW ZEALAND: Andrea Herbert, PhD, University of Canterbury; Daniela Liggett, PhD, University of Canterbury; NIGERIA: Ademola Ajuwon, PhD, University of Ibadan; PAKISTAN: Nisar Ahmed Shar, PhD, CoPI-National Center in Big Data & Cloud Computing; Bilal Ahmed Usmani, PhD, NED University of Engineering and Technology; PERU: Rosario Mercedes Bartolini Martínez, PhD, Instituto de Investigacion Nutricional; Hilary Creed-Kanashiro, M.Phil., Instituto de Investigacion Nutricional; PORTUGAL: Paula Sim?o, MD, S. Pneumologia de Matosinhos; RWANDA: Pierre Claver Rutayisire, PhD, University Rwanda; SAUDI ARABIA: Abu Zeeshan Bari, PhD, Taibah University; SLOVAKIA: Iveta Nagyova, PhD, PJ Safarik University - UPJS; SOUTH AFRICA: Jason Bantjes, PhD, University of Stellenbosch; Brendon Barnes, PhD, University of Johannesburg; Bronwyne Coetzee, PhD, University of Stellenbosch; Ashraf Khagee, PhD, University of Stellenbosch; Tebogo Mothiba, PhD, University of Limpopo; Rizwana Roomaney, PhD, University of Stellenbosch; Leslie Swartz, PhD University of Stellenbosch; SOUTH KOREA: Juhee Cho, PhD, Sungkyunkwan University; Man-gyeong Lee, PhDc, Sungkyunkwan University; SWEDEN: Anne Berman, PhD, Karolinska Institutet; Nouha Saleh Stattin, MD, Karolinska Institutet; SWITZERLAND: Susanne Fischer, PhD, University of Zurich; TAIWAN: Debbie Hu, MD, MSc, Tainan Municipal Hospital; TURKEY: Yasin Kara, MD, Kanuni Sultan Süleyman Training and Research Hospital, Istanbul; Ceprail Şimşek, MD Health Science University; Bilge Üzmezoğlu, MD, University of Health Science; UGANDA: John Bosco Isunju, PhD, Makerere University School of Public Health; James Mugisha, PhD, University of Uganda; UK: Lucie Byrne-Davis, PhD, University of Manchester; Paula Griffiths, PhD, Loughborough University; Joanne Hart, PhD, University of Manchester; Will Johnson, PhD, Loughborough University; Susan Michie, PhD, University College London; Nicola Paine, PhD, Loughborough University; Emily Petherick, PhD, Loughborough University; Lauren Sherar, PhD, Loughborough University; USA: Robert M. Bilder, PhD, ABPP-CN, University of California, Los Angeles; Matthew Burg, PhD, Yale; Susan Czajkowski, PhD, NIH - National Cancer Institute; Ken Freedland, PhD, Washington University; Sherri Sheinfeld Gorin, PhD, University of Michigan; Alison Holman, PhD, University of California, Irvine; Jiyoung Lee, PhD, University of Alabama; Gilberto Lopez ScD, MA, MPH, Arizona State University and University of Rochester Medical Center; Sylvie Naar, PhD, Florida State University; Michele Okun, PhD, University of Colorado, Colorado Springs; Lynda Powell, PhD, Rush University; Sarah Pressman, PhD, University of California, Irvine; Tracey Revenson, PhD, University of New York City; John Ruiz, PhD, University of Arizona; Sudha Sivaram, PhD, NIH, Center for Global Health; Johannes Thrul, PhD, Johns Hopkins; Claudia Trudel-Fitzgerald, PhD, Harvard T.H. Chan School of Public Health; Abehaw Yohannes, PhD, Azusa Pacific University. **Students**: AUSTRALIA: Rhea Navani, BSc, Monash University; Kushnan Ranakombu, PhD, Monash University; BRAZIL: Daisuke Hayashi Neto, Unicamp; CANADA: Tair Ben-Porat, PhD, Tel Aviv University; Anda Dragomir, University of Quebec at Montreal (UQAM) and CIUSSS-NIM; Amandine Gagnon-Hébert, BA, UQAM; Claudia Gemme, MSc, UQAM; Vincent Gosselin Boucher, University of Quebec at Montreal (UQAM) and CIUSSS-NIM; Mahrukh Jamil, Concordia University and CIUSSS-NIM; Lisa Maria Käfer, McGill University; Ariany Marques Vieira, MSc, Concordia University; Tasfia Tasbih, Concordia University and CIUSSS-NIM; Maegan Trottier, University of Lethbridge; Robbie Woods, MSc, Concordia University; Reyhaneh Yousefi, Concordia University and CIUSSS-NIM; FRANCE: Tamila Roslyakova, Université de Montpellier; GERMANY: Lilli Priesterroth, Mainz University; ISRAEL: Shirly Edelstein, Hebrew University-Hadassah School of Public Health; Tanya Goldfrad, Hebrew University-Hadassah School of Public Health; Ruth Snir, Hebrew University-Hadassah School of Public Health; Yifat Uri, Hebrew University-Hadassah School of Public Health; NEW ZEALAND: Mohsen Alyami, University of Auckland; NIGERIA: Comfort Sanuade; SERBIA: Katarina Vojvodic, University of Belgrade. **Community Participants**: CANADA: Olivia Crescenzi; Kyle Warkentin; DENMARK: Katya Grinko; INDIA: Lalita Angne; Jigisha Jain; Nikita Mathur, Syncorp Clinical Research; Anagha Mithe; Sarah Nethan, Community Empowerment Lab.

---

**Supplement Table S2.**
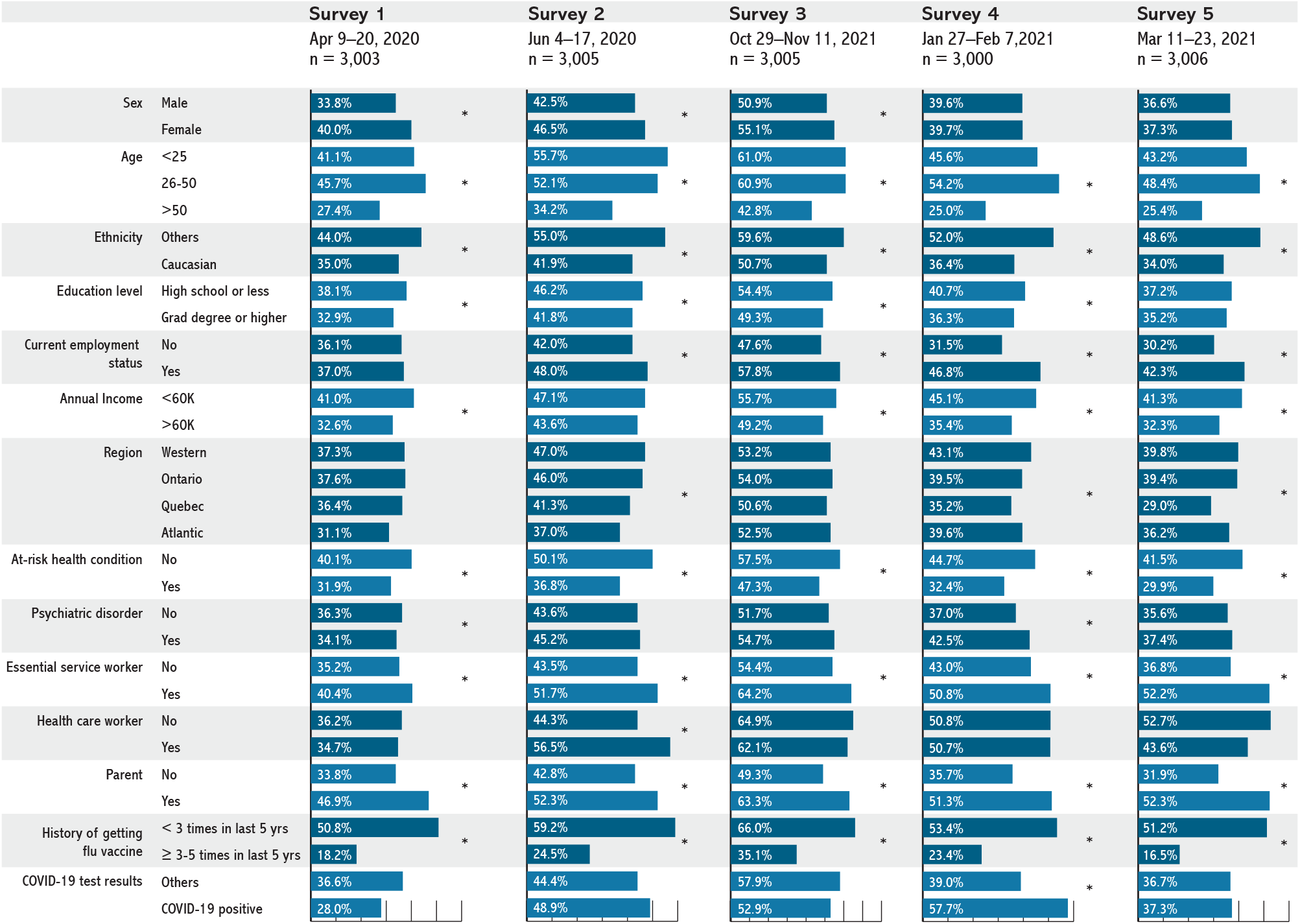
Participant characteristics presented as a function of being hesitant* vs extremely likely to get a COVID-19 vaccine across the three surveys: univariate analyses.

|  |  | Survey 1<br>Apr 9–20, 2020<br>n = 3,003 | Survey 2<br>Jun 4–17, 2020<br>n = 3,005 | Survey 3<br>Oct 29–Nov 11, 2021<br>n = 3,005 | Survey 4<br>Jan 27–Feb 7, 2021<br>n = 3,000 | Survey 5<br>Mar 11–23, 2021<br>n = 3,006 |
| --- | --- | --- | --- | --- | --- | --- |
| Sex | Male | 33.8% | 42.5% | 50.9% | 39.6% | 36.6% |
|  | Female | 40.0% | 46.5% | 55.1% | 39.7% | 37.3% |
| Age | <25 | 41.1% | 55.7% | 61.0% | 45.6% | 43.2% |
|  | 26-50 | 45.7% | 52.1% | 60.9% | 54.2% | 48.4% |
|  | >50 | 27.4% | 34.2% | 42.8% | 25.0% | 25.4% |
| Ethnicity | Others | 44.0% | 55.0% | 59.6% | 52.0% | 48.6% |
|  | Caucasian | 35.0% | 41.9% | 50.7% | 36.4% | 34.0% |
| Education level | High school or less | 38.1% | 46.2% | 54.4% | 40.7% | 37.2% |
|  | Grad degree or higher | 32.9% | 41.8% | 49.3% | 36.3% | 35.2% |
| Current employment status | No | 36.1% | 42.0% | 47.6% | 31.5% | 30.2% |
|  | Yes | 37.0% | 48.0% | 57.8% | 46.8% | 42.3% |
| Annual Income | <60K | 41.0% | 47.1% | 55.7% | 45.1% | 41.3% |
|  | >60K | 32.6% | 43.6% | 49.2% | 35.4% | 32.3% |
| Region | Western | 37.3% | 47.0% | 53.2% | 43.1% | 39.8% |
|  | Ontario | 37.6% | 46.0% | 54.0% | 39.5% | 39.4% |
|  | Quebec | 36.4% | 41.3% | 50.6% | 35.2% | 29.0% |
|  | Atlantic | 31.1% | 37.0% | 52.5% | 39.6% | 36.2% |
| At-risk health condition | No | 40.1% | 50.1% | 57.5% | 44.7% | 41.5% |
|  | Yes | 31.9% | 36.8% | 47.3% | 32.4% | 29.9% |
| Psychiatric disorder | No | 36.3% | 43.6% | 51.7% | 37.0% | 35.6% |
|  | Yes | 34.1% | 45.2% | 54.7% | 42.5% | 37.4% |
| Essential service worker | No | 35.2% | 43.5% | 54.4% | 43.0% | 36.8% |
|  | Yes | 40.4% | 51.7% | 64.2% | 50.8% | 52.2% |
| Health care worker | No | 36.2% | 44.3% | 64.9% | 50.8% | 52.7% |
|  | Yes | 34.7% | 56.5% | 62.1% | 50.7% | 43.6% |
| Parent | No | 33.8% | 42.8% | 49.3% | 35.7% | 31.9% |
|  | Yes | 46.9% | 52.3% | 63.3% | 51.3% | 52.3% |
| History of getting flu vaccine | < 3 times in last 5 yrs | 50.8% | 59.2% | 66.0% | 53.4% | 51.2% |
|  | ≥ 3-5 times in last 5 yrs | 18.2% | 24.5% | 35.1% | 23.4% | 16.5% |
| COVID-19 test results | Others | 36.6% | 44.4% | 57.9% | 39.0% | 36.7% |
|  | COVID-19 positive | 28.0% | 48.9% | 52.9% | 57.7% | 37.3% |

**Supplement Table S1.** Participant characteristics presented as function of survey (weighted proportions)

|  | Survey 1<br>Apr 9–20, 2020<br>n = 3,003 | Survey 2<br>Jun 4–17, 2020<br>n = 3,005 | Survey 3<br>Oct 29–Nov 11, 2021<br>n = 3,005 | Survey 4<br>Jan 27–Feb 7, 2021<br>n = 3,000 | Survey 5<br>Mar 11–23, 2021<br>n = 3,006 |
| --- | --- | --- | --- | --- | --- |
| Variable | N (SD) | N (SD) | N (SD) | N (SD) | N (SD) |
| **Sex** |  |  |  |  |  |
| Male | 1452 (48.5) | 1443 (48.3) | 1442 (48.3) | 1444 (48.3) | 1455 (48.5) |
| Female | 1544 (51.5) | 1545 (51.7) | 1545 (51.7) | 1543 (51.7) | 1546 (51.5) |
| **Age (Numerical)** | 48.0 ± 17.1 | 47.2 ± 17.1 | 48.0 ± 17.1 | 48.0 ± 17.4 | 48.4 ± 17.0 |
| **Age (Categorical)** |  |  |  |  |  |
| ≤ 25 years | 353 (11.9) | 364 (12.3) | 369 (12.4) | 368 (13.0) | 330 (11.1) |
| 26–50 years | 1235 (41.6) | 1252 (42.2) | 1236 (41.6) | 1188 (40.2) | 1226 (41.3) |
| ≥ 51 | 1382 (46.5) | 1348 (45.5) | 1369 (46.0) | 1382 (46.8) | 1415 (47.6) |
| **Race/Ethnicity** |  |  |  |  |  |
| Non-White | 517 (17.5) | 528 (18.0) | 614 (20.8) | 513 (17.6) | 514 (17.3) |
| White | 2442 (82.5) | 2407 (82.0) | 2343 (79.2) | 2403 (82.4) | 2452 (82.7) |
| **Education level** |  |  |  |  |  |
| High school or lower | 2120 (72.2) | 2111 (72.1) | 2126 (72.2) | 2134 (72.3) | 2152 (72.5) |
| Graduate or postgraduate degree | 817 (27.8) | 817 (27.9) | 817 (27.8) | 816 (27.7) | 818 (27.5) |
| **Current employment status** |  |  |  |  |  |
| Unemployed | 1640 (55.7) | 1493 (51.2) | 1428 (47.9) | 1453 (49.2) | 1399 (47.3) |
| Employed | 1304 (44.3) | 1425 (48.8) | 1555 (52.1) | 1498 (50.8) | 1556 (52.7) |
| **Annual household income** |  |  |  |  |  |
| <\$60,000/year | 1309 (49.1) | 1178 (45.4) | 1278 (48.4) | 1316 (49.5) | 1324 (49.1) |
| ≥\$60,000/year | 1360 (50.9) | 1418 (54.6) | 1364 (51.6) | 1341 (50.5) | 1369 (50.9) |
| **Provincial region** |  |  |  |  |  |
| Western<sup>1</sup> | 940 (31.3) | 941 (31.3) | 941 (31.3) | 939 (31.3) | 941 (31.3) |
| Ontario | 1152 (38.4) | 1153 (38.4) | 1153 (38.4) | 1151 (38.4) | 1153 (38.4) |
| Quebec | 705 (23.5) | 705 (23.5) | 705 (23.5) | 704 (23.5) | 705 (23.5) |
| Atlantic<sup>2</sup> | 206 (6.9) | 206 (6.9) | 206 (6.9) | 206 (6.9) | 207 (6.9) |
| **High-risk health condition<sup>3</sup>** |  |  |  |  |  |
| No | 1582 (53.5) | 1703 (57.5) | 1591 (53.7) | 1630 (55.3) | 1686 (56.9) |
| Yes | 1372 (46.5) | 1259 (42.5) | 1372 (46.3) | 1316 (44.7) | 1277 (43.1) |
| **Psychiatric disorder<sup>4</sup>** |  |  |  |  |  |
| No | 2084 (72.4) | 2169 (75.4) | 2160 (74.2) | 2087 (73.2) | 2180 (74.9) |
| Yes | 794 (27.6) | 709 (24.6) | 750 (25.8) | 765 (26.8) | 729 (25.1) |
| **Essential service worker** |  |  |  |  |  |
| No | 2368 (82.3) | 2408 (83.5) | 2534 (86.5) | 2409 (82.7) | 2472 (85.4) |
| Yes | 509 (17.7) | 477 (16.5) | 397 (13.5) | 503 (17.3) | 422 (14.6) |
| **Health care worker** |  |  |  |  |  |
| No | 2748 (95.6) | 2762 (95.7) | 2792 (95.3) | 2796 (96.0) | 2769 (95.7) |
| Yes | 128 (4.4) | 122 (4.3) | 139 (4.7) | 117 (4.0) | 126 (4.3) |
| **Parent of children <18 yrs** |  |  |  |  |  |
| No | 2307 (78.2) | 2303 (78.8) | 2296 (78.2) | 2293 (78.6) | 2289 (78.7) |
| Yes | 643 (21.8) | 618 (21.2) | 640 (21.8) | 624 (21.4) | 620 (21.3) |
| **COVID-19 test results** |  |  |  |  |  |
| Negative | 2953 (99.4) | 2958 (99.6) | 2971 (99.6) | 2913 (98.4) | 2907 (98.1) |
| Positive | 17 (0.6) | 12 (0.4) | 12 (0.4) | 46 (1.6) | 58 (1.9) |
| **History of flu vaccine** |  |  |  |  |  |
| < 3 times in past 5 years | 1671 (56.7) | 1709 (58.4) | 1699 (57.8) | 1581 (54.2) | 1687 (57.8) |
| ≥ 3–5 times in past 5 years | 1275 (43.3) | 1219 (41.6) | 1243 (42.2) | 1336 (45.8) | 1230 (42.2) |

1. **Western provinces:** British Columbia, Alberta, Saskatchewan, and Manitoba 2. **Atlantic provinces:** Nova Scotia, New Brunswick, Prince Edward Island, and Newfoundland/Labrador 3. **High-risk health conditions:** cardiovascular disease, chronic respiratory disease, diabetes, obesity, cancer, autoimmune disease 4. **Psychiatric disorders:** any mood and/or anxiety disorder and dementia

